# Effective clinical caseload management strategies from the perspective of community consultant psychiatrist: Qualitative analysis

**DOI:** 10.1101/2021.05.13.21257049

**Authors:** Nishchint Warikoo, Ezgi Sen Demirdogen

## Abstract

**Objective:** There is a lack of guidance and literature on determining a safe caseload size and how community consultant psychiatrists (CCPs) manage their caseload. This paper therefore aims at exploring effective and safe ways of clinical caseload management by gaining a qualitative understanding of caseload management (CLM) practice of CCPs.

**Design:** Cross sectional Qualitative research using semi structured interviews.

**Setting:** The participants were CCPs working in National health service in Hampshire areas of United Kingdom.

**Participants:** The target population comprised 11 CCPs working in the National Health Service (NHS) to get their view on current practice in NHS and compare past and present practices of CLM.

**Main Outcome Measures:** A qualitative research method was used to explore the topic of CLM by collecting data through observations, interviews, questionnaires and then analysing the data using the coding and emergent themes method.

**Results:** Caseload size for CCPs was higher than a manageable level and had an impact on their ability to service their other responsibilities such as strategic work; they did not have a shared view on setting a limit to their caseload. Majority of CCPs were not using CLM and did not have enough control on limiting their caseload size. Some CCPs were using time management and audit of caseload as effective CLM strategies. NWW was not being used equitably.

**Conclusions:** Although the study represents the perceptions of limited number of CCPs, the findings of this study are unique and an important addition to the slight literature that exists on this topic. The results were in line with existing research that large caseloads can have negative impact on CCPs and their ability to provide effective care to the clients. The key factors determining the caseload size were highlighted. Proactive time management and proactive caseload size management were found to be effective tools for CLM. These supported by job planning meetings, Yearly appraisal, use of electronic data system and New ways of Working could be effective in maintaining a safe caseload size for the provision of safe and effective care. The data from this study can be used for requisite quantitative studies on a larger and statistically significant number of CCPs to find effectiveness of each CLM strategy.

## 1. Introduction

A consultant physician is defined as a senior doctor who practices in one of the medical specialties and accepts ultimate responsibility for the care of patients referred to them and for the clinical work undertaken by other health workers. It is a position of considerable responsibility such as making clinical decisions in difficult, complex, and uncertain circumstances.^1,2^ This paper focuses on CCP because in recent years there has been growing concern about role definition and responsibilities of CCPs as the responsibilities are not well defined and their role may be exploited. This can be used by other clinicians to avoid taking appropriate responsibility.^3^ The recent guideline from the General Medical Council (GMC) “Consultants however must do their best to ensure that arrangements are in place to monitor standards of care and to identify potential or current problems”^4^ also does not help much to clarify this issue and almost blurriness the line between clinical role and managerial role. Besides being unclear the role is evolving and amassing huge amount of responsibilities leading to multiple expectations from the CCPs.^5^

### 1.1. Multiple expectations

CCPs have to manage multiple expectations. Firstly, the patient or the carers frequently demand to be seen by the consultant instead of other clinicians, which sets up an ethical dilemma for CCPs about deciding whether to give in to such requests or to use her/his time in a more efficient way, for example by supervising a group of clinicians.^6,7^ Secondly CCPs are expected to contain anxiety of the team and supporting them in developing a formulation and management plans for complex cases.^5,8^ Thirdly, the clinical record keeping is a responsibility in itself and takes a lot of time particularly when the caseload is large and the risks and the complexity of the cases are high.^3,9^

### 1.2 Stress factors and impact

In addition to the stress caused by the multiple expectation the CCP role has inherent stress factors such as caring for long-term and seriously ill patients, the high demands of dealing with chronic relapsing illness, patients’ suicide risk, fear of violence, heavy workloads and legalistic frameworks - all of which can impact on the CCPs’ own well-being.^10,11^ Many consultant psychiatrists in the UK had started feeling such increased work pressures and gradual erosion in their job satisfaction after integration of consultants into the National Health Service (NHS) of the United Kingdom mainly associated with increased case-loads and high stress levels. The wide range of work stress can cause emotional exhaustion, severe depression and severe consequences such as suicide and retirement.^6,12-,16^ Furthermore, these also have unfavorable impact on effectiveness of CCPs and the service being provided to individual clients which may lead to significant public health consequences in terms of morbidity and mortality.^14,15^

### 1.3 Current guidelines

The issues mentioned earlier are not new and have been acknowledged two decades ago leading to the government initiative The New Ways of Working (NWW) which supported low caseloads and limited responsibility for a CCP.^17^ NWW did not however provide specific guidelines on the number of cases that the consultant should hold.

Unfortunately, this specific issue of caseload size has not received much attention from the researchers in the past; there is no study that deals with the issue of CLM by the CCPs’ and there are no guidelines for CCPs to be able to objectively assess and manage their caseloads. Considering the deficiency in the literature on this subject a qualitative research on this topic of CLM, focusing on the CCPs was planned with aim of exploring and examining more effective and safer ways of CLM which could help reduce pressure on CCPs in particular and the service thereof. This could be achieved by gathering views of CCPs on management of large caseloads - gain insight into factors influencing the caseload size, individual clinician’s decision-making process and methods used to set a limit to their caseload and to identify effective evidence-based tools used in caseload size management. As there is not enough research evidencing the progress, implementation or effectiveness of NWW this study could also generate data on effect of NWW on the day-to-day clinical work of a CCP

## 2. Method

### 2.1 Study Design and Procedure

Primary qualitative research method was used in order to get a broad view and to explore in detail the complex topic, CLM by the CCPs and to generate hypothesis and questions. Qualitative approach helped by doing a survey of primary data i.e. knowledge, experience, expectations, or preference of the CCP group and enabled the study of problems and the issues in their natural setting with better understanding of the complex interplay of any variables and generating hypothesis. The fact that there is no hypothesis has guided the nature of the study and the design of this project followed a logical sequence enabling each step to act as a building block for the next step (Supplementary Fig 1):

1. **Researcher**’**s experience and reflections:** The researcher started with reflection on his own clinical experience (CE), which acted as a guide to the literature search (LS).
2. **The literature review** was conducted within a sound structural concept with careful searches to find relevant data and information. This generated themes which then underpinned the development of the structure for the semi-structured interview.
3. **Semi structured interview** (ninety minutes long with a senior CCP) In view of the lack of literature from which to draw conclusions, the researcher had to use primary data gained through semi-structured interview and questionnaire. The themes generated from CE and LS formed the basis of a semi-structured interview (SSI) which was conducted with one CCP.
4. **Semi-structured questionnaire (SSQ) (Appendix):** These previous steps generated key themes-: Multiple responsibilities, large caseloads, caseload size is dependent on complex interactions, personality factors, effects of large caseload, training for CLM, CLM approach, NWW. These emerging themes from the CE, LS and SSI were used to develop a Semi-structured questionnaire (SSQ) using the Likert Questions to help ascertain how strongly the CCPs feel about the statement. Open-ended questions were used to allow the expression of opinions in a free-flowing manner and multiple-choice questions where respondents were restricted to choose among any of the given multiple-choice answers. The 11 CCPs working in child and adolescent psychiatry in the Hampshire area of England, who had expressed their interest, following a presentation by researcher about the aim of the project were included in the study. Fully informed consent was obtained voluntarily from each participant before they were given the SSQs. Ten CCPs completed and returned the SSQ.
5. **Post questionnaire interview:** Four CCPs who had completed the SSQ were interviewed about the SSQ quality to support the validity of the questionnaire.

The data generated from LS, SSI and the SSQ were then analysed using the coding and emergent themes method. To improve the validity of qualitative method of the study, more than one method was used in combination.18 In addition to SSI and SSQ, follow up interviews were used to gather evidence on the validation of the SSQ. The participants were asked to do the SSQ in their own time which minimised any effect that presence of researcher might have on the data gathering.^18^ Coding and thematic analysis was used to help present the information in an easily readable form.

All procedures of this study were approved by Clinical Ethics Committee of Sussex partnership NHS foundation trust. All procedures performed in this study was conducted in accordance with the Declaration of Helsinki regarding research on Human participants. The researcher was aware of the ethical issues due to his role as a researcher observing the same set up in which he was working and has considered this in the analysis, conclusions, and recommendations. Importantly the researcher was well placed to do this research as it would have been “inconceivable that such interview could have been conducted by someone with no views at all and no ideological or cultural perspective”.^18^

### 2.2 Participants

In order to gain an in-depth understanding of the experience of the CCP, a group from the child and adolescent psychiatry was selected as literature review suggested similar challenges are faced by the CCP from different sub-specialties of psychiatry.^7,19^ Focusing on one sub specialty as a representative of the psychiatrist as a wider group avoided the masking of similar issues representative of all sub specialties by the subtle differences between the sub specialties. It also enabled seeking out the individual and groups who fit the bill instead of having an average view of the population.^7,19^

The target population comprised 11 CCPs who were currently working with the NHS so that they can give their view about current practice in NHS or compare past and present practices of CLM. The participants were not placed in separate groups since the comparison was not the aim of the project. The number of participants was decided pragmatically rather than by a formal calculation. On balance an in-depth interview with one experienced CCP and 11 SSQ with 11 CCPs was considered a reasonably practical sample size to generate enough views and the right number for this type of research. Extensive and long career of the experienced CCP in psychiatry, her experience of working in both traditional way as a CCP and with the NWW and her multiple roles, both clinical and managerial was in concordance with aim of the study

Consultants working in inpatient units were excluded from the research as the finite number of available hospital beds means there is a structural cap on their caseloads. The CCPs who have been in lead roles for implementation of NWW were also excluded as they may have a more favourable view of the NWW. The CCPs working in the same mental health team as the researcher were also excluded to avoid any undue influence when recruiting for the project.

### 2.3 Data analyses

Coding and thematic analysis was used to capture the richness of data from each step and present it in an easily readable form. Thematic analysis was conducted according to the guidelines presented by Jennifer (2001).^20^ The codes in the coding framework had quite explicit boundaries (definitions) assuring that they are not interchangeable or redundant and enabling limited scope and focused explicitly on the object of analysis. Coding the textual data was based on the theoretical interests guiding the research aim and based on salient issues that arose in the text; thus, preventing the coding of every sentence in the original text. Following this crucial step of detecting patterns in the text, the codes were used to dissect the textual data into meaningful and manageable text segments.^20^ Repeated review of text segments in each code and group of related codes enabled identification of underlying patterns and structures and led to emergence of common and significant themes (Outcome themes). Then outcome themes were refined further into discrete (non-repetitive) themes encompassing a set of ideas contained in numerous text segments, which shaped the data into a manageable set of significant themes (Table 4).

## 3 Results

### 3.1 Results of the literature review

Literature review gave an overview of relevant literature and research to date hence forms the background to this research project. The first literature search generated 38 papers which did not cover all the relevant issue of CLM by CCPs. The second literature search was conducted on the papers focused on cause of the stress and burnout and some suggestions of how to prevent this, however none of the papers were on the CLM by CCPs. The third literature search generated 26 papers some of which were about CLM by case managers however only one paper was for CCPs.^21^ Solutions from literature review were explored and discussed under two headings; wider system level solutions and individual CCP level solution.

#### 3.1.1 Change in the System - Introduction of New Ways of Working (NWW)

The literature review highlighted NWW as one of the key solutions for reducing caseload and responsibility on the CCPs.^17,22^ NWW encouraged the CCPs and the team to support the CCP to have a small caseload so that the time saved can be used by CCP to provide consultation to the team. NWW recommended that consultant psychiatrists should not have a large caseload of patients and the responsibility should be distributed amongst team members rather than delegated by a single professional, such as the consultant.^22^ NWW encouraged the multidisciplinary teams to operate dispersed leadership based on appropriateness of capabilities of team members. In theory all the above measures could be effective in reducing two stressors on the CCP (i.e. large caseload and associated responsibility). The CCP is in turn expected to take on other responsibilities with the time that could be freed up with having a smaller caseload; supervising and supporting staff, learning and development, research, service improvement, etc. and responding in a flexible and timely manner as needed to help manage the care of those with more complex needs.^23^ This in theory would limit the responsibility of the CCP and making appropriate and effective use of their time. However, there were some limitations to NWW highlighted by the report, particularly the possible role erosion that a consultant might feel due to distributed responsibility. The other caution was the need for additional resources in the team to provide support for the new roles of the clinicians including training them to take on new roles. ^17,22,23^ The other disadvantage of a purely consultative role might be that constantly dealing with such high stress cases can be destructive to the clinician”.^2^ Furthermore, there would be financial implication of NWW as the change in the consultant role would need team support and up skilling of other clinicians to take on new and varied roles. However, literature search did not generate any further progress document on NWW or a recent update on the NWW’s implementation or effectiveness.

#### 3.1.2 Solutions at the CCP level

1. **Training in leadership:** The NWW proposed a cultural shift in the way the service is provided and meant a CCP has to have a smaller caseload, be very boundaried in accepting new cases and to enable the team members to take on the newly developed roles and responsibilities.^4^ The report on new roles for psychiatrists reporting the findings of two consultation days attended by over 600 delegates in 2003, recommended the need for training in leadership and management (BMA,2004; published by the British Medical Association and authored by the National working group).^2^ The report highlighted that training of psychiatrists would enable them to gain the ability to be a team leader and understanding of group dynamics within teams so that they could avoid accepting covert responsibility for the convenience of others rather than where appropriate.^2^ In addition to the training there is a need for tangible evidence to support their decisions on caseload number and their use of clinical time.
2. **Audit and regular review of the caseload:** The paper by King (2009) based on a survey of 188 case managers used an online cross-sectional survey with both purpose-developed items investigating methods of case allocation and caseload monitoring and standard measures of work-related stress and case manager personal efficacy.^24^ The paper highlighted that the higher caseloads were associated with higher level of work-related stress and lower level of case manager personal efficacy. King found that regardless of the caseload size “active monitoring of caseload was associated with lower scores for work-related stress and higher scores for case manager personal efficacy”. These results around the active monitoring of caseload were promising.
3. **Need for time management:** Watson (1985) suggested that service couldn’t’t be assumed to be better simply because more people are providing it.^25^ Creed (1995) suggested that consultants should review their out-patient clinics to establish their precise purpose and plan packages of care (in terms of time and treatment skills required) that allow an estimate of the resources spent on each patient.^7^ Creed (1995) highlighted the need for consultants to audit their time in order to review their way of working, increase their efficiency and be able to inform their managerial colleagues of this aspect of a quality service.^7^ The benefit of time management as a stress-reducing strategy was also evidenced in a survey of 37 consultant psychiatrists in 2007.^9^ On similar lines in 2004, a new contract for consultants was introduced with inbuilt mechanism for consultants to complete a diary exercise. This diary was to be used as the basis of negotiations for any changes in job plan. The aims of the contract were to properly quantify and reward the work of the consultants.^6^
4. **Using a consultation model in child and adolescent mental health services:** The paper most relevant to CLM ^21^ was designed to survey a different model of consultant role in a child and adolescent mental health service to address the problem of an on-going waiting list. The method used was a log of referrals was kept from date of entry and when patients were seen. The main outcome was in reducing the waiting time from 22 weeks to 8.4 weeks. The conclusion was that the consultants should consider using a consultation model in child and adolescent mental health services. Though there were weaknesses in the model and design of the study, it was probably the only study which studies a model similar to that proposed by NWW.^17,22,23^ The learning points were that the purely consultative model could work, given the other variables were favourable, e.g. the skill mix and number of clinicians in the team.

### 3.2 Results of Semi structured interview with a CCP 1- themes

The senior CCP with whom SSI was conducted defined “limiting caseload” as a complex topic because of its dependence on the number of other clinicians in the team and their skill mix highlighting the impossibility of having a consultation and strategic role while holding large caseload. CCP found it difficult to take on the NWW, possibly due to individual personality characteristic which was found to be similar according the CCP when CCPs were working in a traditional system where large caseloads led to clinician anxiety and “often cases were lost to follow up”. According to her, traditional system had its advantages as the assessments were comprehensive and she found greater satisfaction in her work and NWW could make clinicians feel they were not able to use all their clinical skill and they might not get enough job satisfaction. However, she acknowledged traditional style of working was not sustainable as it did not leave much spare time to provide consultation and agreed there was a need for CLM but was not aware of a CLM tool. She agreed with the view that CCPs might benefit from managing their caseload and time management. This she feels can only be done by proper support from the team.

CCP 1 (2014) highlighted the need of each doctor to have a yearly appraisal. In which the doctors would discuss their practice and performance with their appraiser and use supporting information to demonstrate that they were continuing to meet the principles and values set out in good medical practice.^26^ She highlighted that the preparation for appraisal and the appraisal itself could provide an opportunity for a CCP to reflect on their caseload and the use of clinical time which also points out the need for the CCPs to continue professional development (CPD) and to ring fence the time for CPD.

### 3.3. Results of the semi structured questionnaires

The SSQ (see appendix) was completed with 90 % response rate.

#### 3.3.1 Caseload size

Majority of CCPs had high caseload; 80 percent of CCPs had more than 100 cases, 60 percent had more than 150 cases each. Even with the introduction of the NWW and the time scale since implementation there appeared to be no major reduction in the caseloads. All agreed that it was not possible to carry large caseloads and do strategic work as well. Majority of the CCPs thought they had more cases than the non-medical colleagues in their teams.

#### 3.3.2 Consequences of large caseloads

The adverse impact of high caseloads was felt by majority of CCPs with 80 percent agreeing that bigger caseload does not allow for any consultation time, or any time to provide urgent assessment and eats into their protected continuing professional development (CPD) time (Table 1).

**Table 1-.**
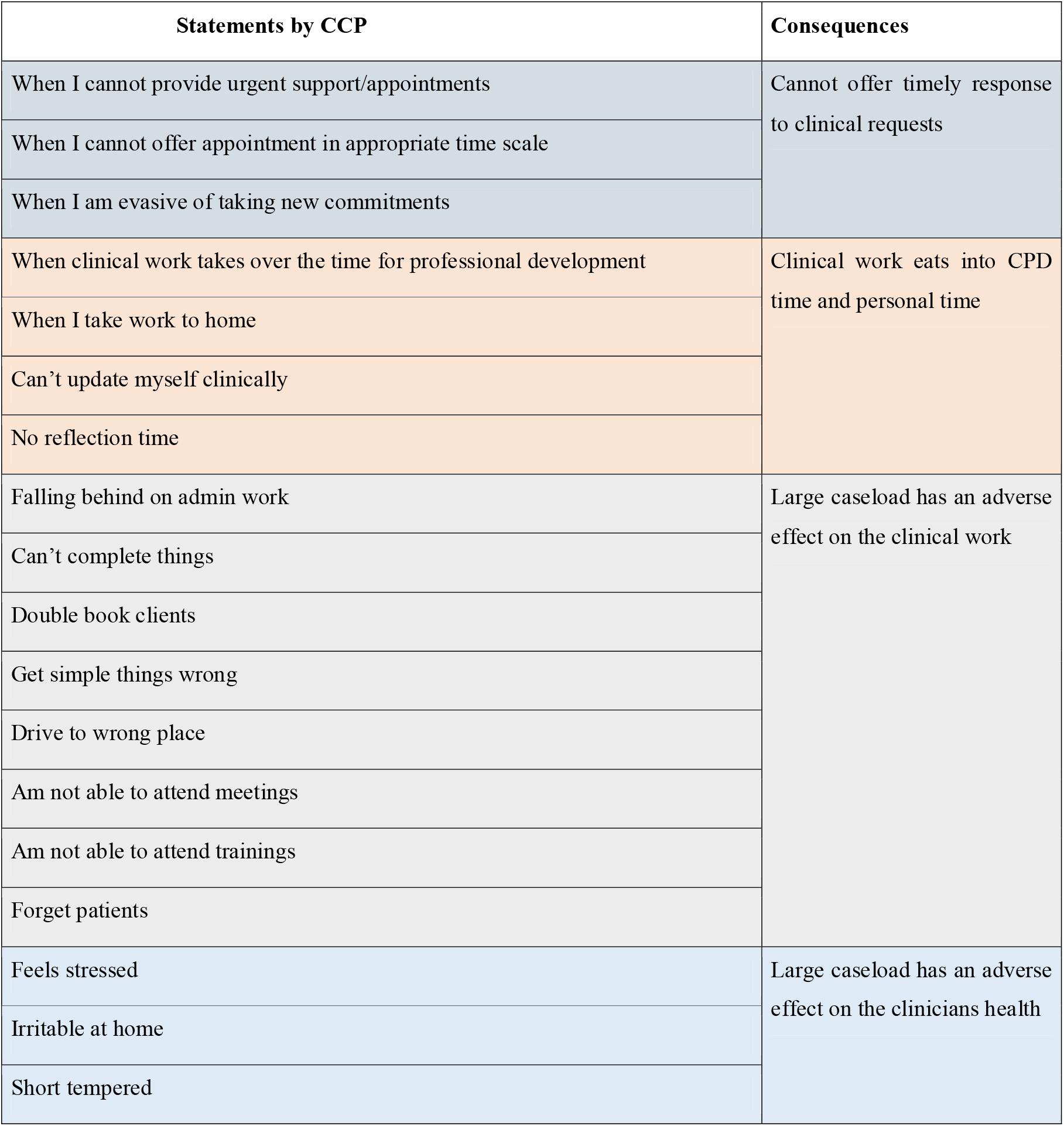
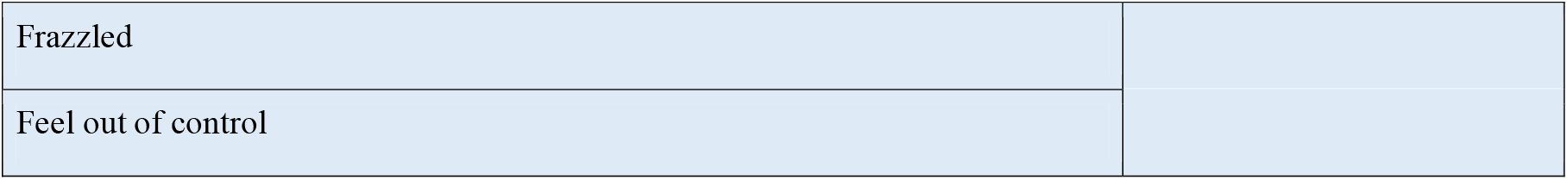
Indicators for large caseload and consequences

#### 3.3.3 CLM

Need for active management of caseload was highlighted with all the CCPs (9 SA & 1A) agreeing that CLM is an important part of the clinic practice. Majority (9-Y and 1-NS) felt the need to proactively manage the caseload however, only 70 percent agreed that each clinician should audit his/her time in order to review their way of working and increase their clinical efficiency. None of the CCPs had decided on the limit of their caseload size before starting their current job (Table 3).

**Table 2-.** Comparison of themes

| Themes from background issues | Themes from SSI | Themes from the questionnaire |
| --- | --- | --- |
| Unsustainable caseloads | Caseload | Large caseloads |
| Multiple responsibilities | Traditional system | Multiple responsibilities |
| Changing roles | Stress |  |
| Collaborative working |  | Caseload size is dependent on complex interactions |
| Role specificity | Personality characteristics affect case load size |  |
| Impact on health | Effect of large caseloads |  |
| Consequences | Consequences | Effects of large caseload |
|  | No guidelines |  |
| **Themes from Literature review** | NWW has not been taken up | Personality factors |
| NWW | Benefits of NWW | NWW |
| CCP to audit caseload | Audit case load and manage time | CLM |
| CCP to manage clinical time |  |  |
| Formal training required | Training needed for CLM | Training for CLM |
|  | Future of NWW |  |
|  | CLM future support | Effective CLM tool Strategy |

**Table 3-.** How do CCPs decide on the size of caseload: Strategies used by CCPs

| Statements of the CCPs' | Theme |
| --- | --- |
| I don't have a formula to decide | No decision |
| I don't decide |  |
| I respond to service needs | Passively Respond |
| It is driven by the needs of the service |  |
| Driven by the capacity of other psychiatrist in the team. |  |
| Work later in the evening | Overwork |
| Work late at home |  |
| Don't see patients that frequently | Unfavourable quality of work |
| See too many patients a day |  |
| Plan own diary | Proactively manage caseload |
| Keep space for CPD, admin and management work |  |
| Define the number of patients I see in one week |  |
| Try not to caseload | Proactive offloading of cases |
| Review caseload regularly |  |
| Identify cases to relocate |  |
| Consider early discharge |  |

CCPs expressed a sense of passivity with majority (80 percent) agreeing that their caseload was not under their control but was determined by the service needs. About 70 percent thought their caseload size and complexity was dependent on the skill mix of other clinicians in the team and similar numbers felt that their caseload size and complexity was dependent on the ratio of the CCPs to non-medical clinicians in the team. (Table 3).

Some CCPs managed their work by cutting corners “short letters” and seeing patients less often and spending less time with the patients. Some CCP used proactive approach to number of scheduled patient contacts; some of them were actively deciding the number of patients they see each week, leaving space for consultations and for any urgent requests whereas some of them proactively managed their caseloads by limiting the number of new referrals they take, proactively discharging cases, referring to others clinicians and sign posting cases. Some CPPs also used a pragmatic approach to use of available time; they planned their diary proactively and blocked time slots for their CPD, admin work and management time. Though some CCP’s had success with this active CLM; one CCP expressed his/her inability “I make a list of the cases but couldn’t follow it through; am too busy.” Some clinicians supported their active CLM by referring to their job plan and seeking support from the multidisciplinary team. They do however highlight the lack of guidelines. Not even one CCP could provide information about any guidelines on the CLM (Table 4).

**Table 4-.** CCPs manage their caseload by doing the following:

| Statements of the CCPs' | Themes |
| --- | --- |
| Don't manage | Passive approach |
| Deal with what comes |  |
| Use CPD time for admin work | Overwork and use personal time to complete clinical work |
| Work at home |  |
| Late evenings |  |
| Short letters | Cut corners to manage caseload |
| Minimise contact |  |
| Short appointments |  |
| Allocate as soon as possible | Reduce caseload |
| Discharge to GP |  |
| Close piece of work |  |
| Cases to clinics |  |
| Involve team for queries |  |
| Refer cases to care coordinators |  |
| Encourage team to refer complex cases only | Reduce intake |
| Consult other clinicians |  |
| Support others and don't take new cases |  |
| Review with CCP |  |
| Try not to case hold |  |
| Refer to job plan | Structure to help maintain boundaries |
| Keep record |  |

#### 3.3.4 NWW

The take up for NWW was low with only about 25 percent CCPs actively using NWW and 20 percent not feeling certain whether NWW was being used in the team. One CCP noted some positive impact of NWW on his/her clinical work but the evidence was not enough to determine the effectiveness of NWW (Supplementary Fig 2).

## 4. Discussion

This study was set out to explore the views of CCPs on caseload sizes, gain insight into their current CLM practice and the factors influencing this process. Our results provided information about the factors influencing the size of caseload and generated some data to highlight the effectiveness of active case management approaches which can help CCPs be more effective care providers. The analysis of participants ‘opinions and comments revealed that CCPs had similar perspectives on these topics. It indicated current caseload size does not allow enough time to provide urgent assessment or for consultation and eats into their protected CPD time hence impinging on provision of effective care. Findings also highlighted CCPs’ did not have enough control on their own caseload size particularly because they were not using CLM and implementation of NWW was not good enough to stop large caseload thus leading to negative consequences.

### 4.1 Caseload size and possible determinant factors

Gathering views of CCPs (Table1) on management of large caseloads helped us to gain some insight into the individual clinician’s decision-making process around CLM and generated information about the factors that can influence the size of caseload which was higher than a manageable level.

This persistence and pervasiveness of high caseloads can be understood by taking a historical perspective starting at CCPs training years when they start holding large caseloads. They possibly learnt from some of their supervisors who considered large caseloads as a “badge of honour” or possibly because large caseloads were the norm (researcher’s experience) during training years. In addition, the CCPs in their training programme did not have training of CLM techniques neither were there any guidelines on caseload size. This continued in their CCP posts with no objective guidance on caseload limit. As per the CCPs their job adverts did not suggest a limit to the cases and they started their post with not even a rough idea of how many cases they will be responsible for and alarmingly would not even know when their case-load became too large.

Following on in the CCPs role they start feeling ultimately responsible for the service hence not only are they taking on more and more cases but making allowances for the lack of resources in the team. They thus take on “covert responsibility for the convenience of others rather than where appropriate”.^2^ In line with results from available literature and their own view the CCPs continue to see more cases rather than be more available to the team and continue to have more cases than their non-medic colleagues.^27^ The CCPs also report that as the caseload increases, many CCPs are not able to decide when to stop taking on more cases instead they feel the need to respond to service needs, feel they need to take on cases particularly if other psychiatric colleges are not available or able to take on cases.

There seems to be an absence of an authority to support CCPs decision of not taking on any new cases and too many responsibilities. The NWW was specifically meant to provide guidance to trusts to support CCPs in having low caseloads.^27^ This has not been effective as the CCPs responses show NWW has not been implemented in an even and balanced way. Lastly, CCPs own personality and appetite for risk also seem to have influence the size and management of their caseloads. Different CCPs due to their differing personalities might respond differently to similar work pressures, service needs and available support structures. ^13,23^ As there is no expectation for them to have supervision they do not have any tailored oversight or guidance to guide them with the size of their caseload. Some CCPs are so used to having large caseloads that having a small caseload makes them think they are not doing enough, “I am not pulling my weight”. This is even when the CCPs have the reassurance from the team that their input in form of consultation is more valued as compared to seeing more cases. Some CCPs get a feeling of triumph with large caseload. Fifty percent of CCPs agreed to the statement - some of the CCP’s find holding a large caseload as a “badge of honour”. CCPs with such attitude towards large caseloads might not actively try to reduce their caseload.

### 4.2 Consequences of large caseloads

The data from SSI and SSQ brought up alarming facts (Table1). The CCPs report that as the caseload increases, many CCPs are not able to decide when to stop taking on more cases. They continue to see more and more cases, spend less time with the patients, write brief letters and see the client less often.

Some CCPs, to keep on top of their work, start working longer hours, use up their CPD time and even work in their own time, often at home. As they use up their CPD time there is no time left for reflection, to audit their caseloads or to plan their clinical time effectively. This appears to be a vicious circle where many CCPs are too busy to take out time from their busy schedule to save time.

Majority of CCPs (9 out of 10) strongly agreed with the statement that high workload and work stress can affect the health of a CCP in line with evidence from previous studies. Continuing to work with large caseloads and trying to manage by overwork and using personal time to keep on top of their work can lead to burn out. Higher levels of workload and associated stress have adverse impact on psychiatrist’s health status including serious contemplations like suicide and retirement”.^12,14,23,28^ These results highlight the importance of protecting CCPs health for effective service provision as CCPs central role for the provision of care to patients by direct work with patients or through supervision and consultation to other clinicians.

### 4.3 Effective CLM

Even though many CCPs are not using CLM, the few CCPs who have used it, their approach appears to have worked well for their clinical practice (Table 3, 4). It uses two key principles, management of time and caseload. This approach is consistent with the proposals in the literature review. This approach to CLM uses effective time management consistent with the suggested approach by ^6^ and regular Audit/review of case load which is similar to approach suggested by King (2009).^24^

#### 4.3.1 Time Management

CCPs plan their diary proactively and block time slots for their clinical work, CPD, admin work and management time. This helps decide the available clinical time to see clients in a working week. They then decide on the number of scheduled (planned) cases they can see each day with appropriate time for new cases and lesser time for review cases. There is also some time left for unscheduled work (urgent and emergency cases). Based on this plan the CCPs then decide the number of clients they can see weekly and have some idea of the number of clients they can see over a month. This number is then shared with the team manager and helps in more informed allocation of cases to the CCP.

#### 4.3.2 Caseload management

##### 4.3.2.1. Pragmatic gatekeeping of caseload

One CCP encourages the team to refer only complex cases. This would make best use of his experience, specialism and extensive training; rather than using CCP time for routine cases.

One CCP mainly provides consultation to other clinicians supports other clinicians in their management plan and does not take any new cases. This is an effective way of using CCPs expertise to provide effective care through their care coordinators.

One CCP reviews cases only with a care coordinator and involves the team for queries. This again means the patient will get the consultant input as needed and not by default. They will also get the on-going support from the care-coordinator and from the other clinicians in the team

##### 4.3.2.2. Pragmatic case load shedding

CCPs review case load regularly and identify cases which can be relocated to other clinicians or can be considered for early discharge. Some CCPs refer cases to care coordinators and allocate to clinicians as soon as appropriate, thus creating space for more cases.

Another approach used is to discharge cases to the general practitioner if the input being provided can be safely delegated to the GP.

This CLM model is being used by a limited number of CCPs from this small group of CCPs from a sub specialty of Psychiatry. This is a limitation to the use of this model.

The use of this model provides evidence that an effective model of CLM could help the CCPs manage their stress, possibly “irrespective of the size of their caseload”.^24^ This model, however, needs to be supported by job plans which formalise the agreement on the size of caseloads. The support can also be generated if there are specific guidelines from the Employing Trust, GMC or Royal College of Psychiatrists.

#### 4.3.3 Other support factors for CLM

##### 4.3.3.1 Yearly appraisal

This can ensure that there is a yearly review of the work by the CCP. This will also hopefully mean that CCPs will do a more regular review of their caseload and ring-fence some time for their CPD.

##### 4.3.3.2 The electronic data system (as suggested by CCP1)

The system provides an overview of the caseloads of each clinician and hence will provide a dash board for the caseload of the team. As many clinicians have found it cumbersome to maintain an excel sheet to monitor their caseload, the electronic system may provide an easy option.

##### 4.3.3.3 Job Plan meetings

These are usually held yearly and can enable a CCP to discuss their caseloads and effective use of their time with the clinical lead of the NHS trust and use this as an authority to support their decision on caseload size.

The proactive time management including diary planning, defining time for CPD, admin work and management work helped CCPs plan and protect their clinical and CPD time respectively. Proactive case management including regular audit of caseloads, identifying the cases to be signposted, reallocated and discharged on a regular basis helped maintain a manageable caseload size. The Key factors of the CLM evidenced by this study are effective management of time consistent with the suggested approach by Creed, 1995 and Harrison, 2007 and regular Audit/review of case load which is similar to the approach suggested by King 2009.^6,7,24^ Other CCPs considered using CLM but were unsuccessful possibly for two reasons 1. Initiation - CCPs were very busy and could not allocate time for planning their diary or audit their caseload. 2. Persistence - CCPs started the process but could not persist with the process of regularly reviewing their caseload and planning their diary.

### 4.4 New ways of Working -NWW

The literature review suggested NWW as a comprehensive answer to many issues around the large caseload and associated stress on CCP.^22^ NWW, however, has not been implemented evenly evidenced by views from participants in this study who were from the same NHS Trust, working in the same subspecialty in the same geographical area. With only about 25 percent CCPs actively using NWW suggest a patchy implementation of NWW and suggests that the implementation of NWW was dependent on each CCP and/or their individual team. There are many possible reasons for the poor uptake of NWW including. Firstly, the lack of communication about the NWW to the end user which appears to have hindered implementation of NWW. This view is supported by the results from the questionnaire as many CCPs were not even aware of NWW being implemented in the trust. Secondly, for NWW to succeed the team members need upskilling to take on new and varied roles. This process needs support from the senior management due to the financial implications. This support for various reasons was not present to the teams. Thirdly, the skill mix of the team and the number of other clinicians in the team are other important factors to make implementation NWW possible. For instance, if there is one CCP and one other clinician implementation of NWW will not be possible. Fourthly, CCPs and individual teams ‘appetite for change effects implementation of NWW.

CCPs without adequate supervision may in the changeover to the new system (NWW) feel more anxious and find it daunting to “work indirectly with clients through supervision to other clinicians.” Besides the implementation problems, sustenance of NWW depends on CCPs to give up some traditionally held roles which might generate feeling of loss of professional identity and role erosion which some of the CCPs are already feeling.^2,22^ Decrease of job satisfaction with the shift to NWW is another important consequence. This also indicates that NWW on its own still is not enough to cater for the complexity of the problem and the additional support needed may be in the form of a good CLM method. The actual impact of NWW and extent of its implementation need to be studied in a study focused on NWW.

## 5 Limitations of study

The findings from this study should be interpreted cautiously due to the small sample size. The study was limited to CCPs from one NHS trust and one geographical area and may not be a full reflection of the practices in other areas. The third limitation is the cross-sectional study design which does not allow to study prospectively the changes that would occur over period of time and the direction of the effects among the variables could not be identified. However, this study is a step towards understanding the factors which affect caseload size, since it is the first study to focus on caseload management.

## 6 Conclusions

Despite the limitations, the present study highlights the key factors determining the caseload size and effectiveness of some CLM techniques which can help in management of caseload size as the large caseloads can have negative impact on providing effective care to the clients. The Key factors of the CLM evidenced by this study are effective management of time and regular Audit/review of case load.

The current implementation level of NWW in absence of any CLM techniques are not effective in reducing CCPs caseload and associated stressors.^14,15^ CLM can be initiated and managed by the CCPs giving them control over their caseloads. In addition, steps need to be taken through a concerted, comprehensive and persistent effort at individual CCP, team and organizational level to implement evidence-based solutions like NWW.

Although the study represents a pilot into the perceptions of CCPs working in C&A Psychiatry in a public health organisation, the findings of this study are important and unique; adding to the small amount of literature that already exists on this topic. These findings can underpin a quantitative study to identify effective evidence based CLM techniques.

### What is already known on this topic

Managing large caseload, complexity of the cases and associated risk can be significant stress factors for a community consultant psychiatrist (CCP) which in turn can have an effect on their health and the quality-of-care CCPs provide to the patients.

### What this study adds

CCPsdo not have enough control on their own caseload size because they were not using CLM and have large caseload thus leading to negative consequences for them and quality of care.

Current caseload size does not allow enough time for CCPs to provide urgent assessment or for consultation and eats into their protected CPD time hence impinging on provision of effective care.

Individual clinicians use of Time management and proactive caseload management can be effective tools in CLM and if supported by job planning, appraisals and NWW can provide a robust CLM support structure for the clinician.

## Supporting information

Semistructured Questionnaires

## Data Availability

The data used in this study is available from the researchers on request

## Footnotes

### Contributors

N.W. conceptualised the study, designed the model, conducted the data analyses; E.S.D. aided in interpretation. N.W. and E.S.D wrote the article, reviewed and revised the report. Each author contributed important intellectual content during manuscript drafting or revision and accepts accountability for the overall work by ensuring that questions pertaining to the accuracy or integrity of any portion of the work are appropriately investigated and resolved. All authors approved the final version of the report. The corresponding author attests that all the listed authors meet the authorship criteria and that no others meeting the criteria have been omitted. NW is the guarantor.

### Ethical approval

This research project was reviewed and approved by the Institutional Review Board of the Department of Veterans Affairs Saint Louis Health Care System.

### Data sharing

Authors ‘access to the study data was permitted via a data access agreement. The data associated with the manuscript is not currently publicly available.

The lead author (the manuscript’s guarantor) affirms that the manuscript is an honest, accurate, and transparent account of the study being reported; that no important aspects of the study have been omitted; and that any discrepancies from the study as planned (and, if relevant, registered) have been explained.

### Provenance and peer review

Not commissioned; externally peer reviewed.

### Dissemination to participants and related patient and public communities

There are no plans to disseminate the results of the research to study participants. Study results will be shared with the public via press release, social media, and conference presentations.

---

Results from the semi structured questionnaires

**Figure 1-.**
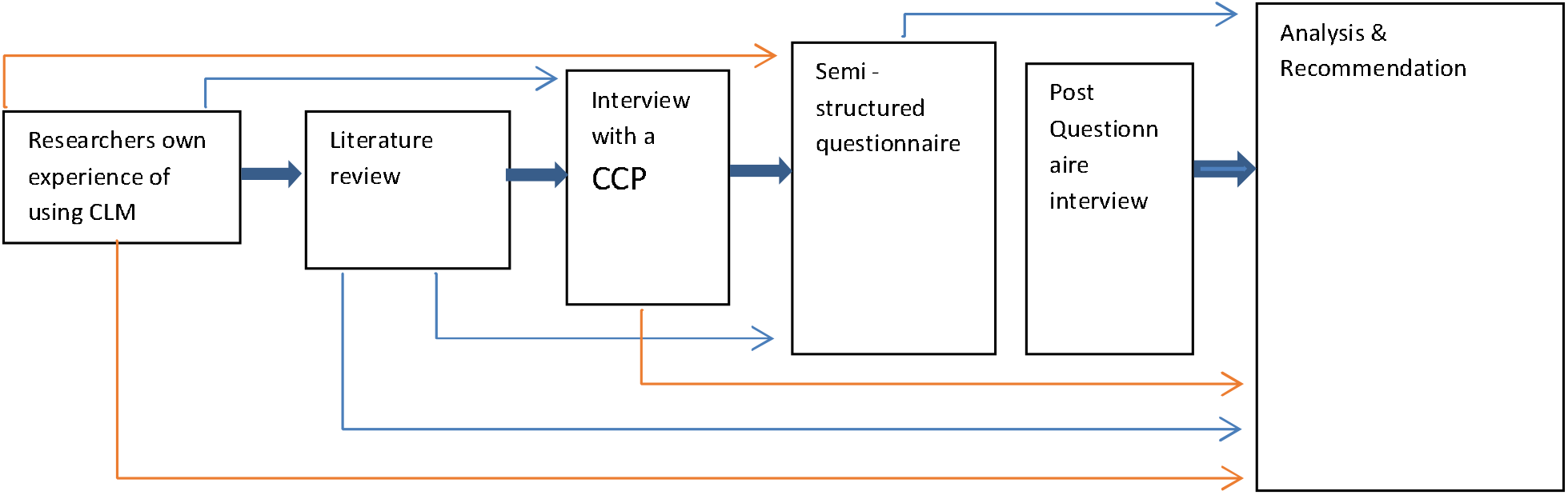
Design of the project.

**Figure 2 -.**
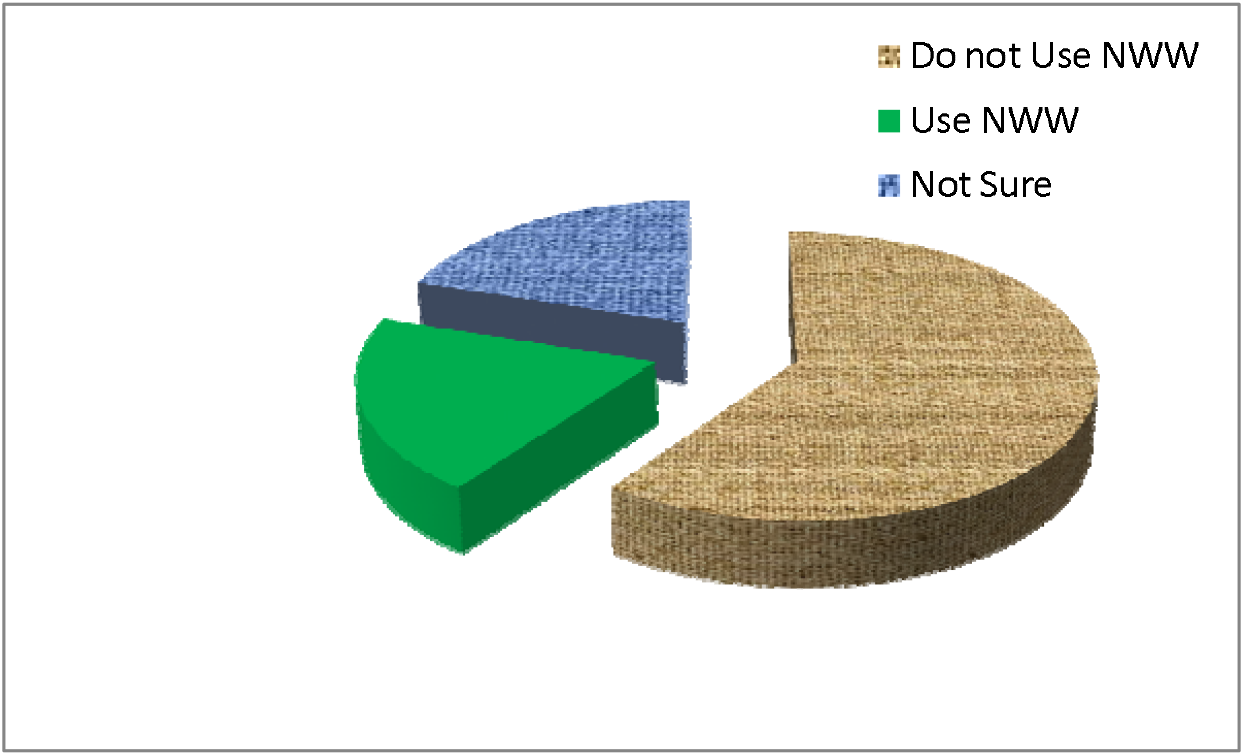
Implementation of NWW.

