## Supplementary material for "Effective clinical caseload management strategies from the perspective of community consultant psychiatrist: Qualitative analysis": Semistructured Questionnaires

**Appendix: Semi Structured Questionnaire**

The perspective of Community Consultant Psychiatrist (CCP) on the case management of large case loads.

The questions and statements in this questionnaire are divided into 4 subsections and are based on Published research and interview with a senior CCP. Please mark your preferred answer by clicking on the box ☐ and it should change to ☒. If it does not change please place an X next to your preferred Box. Please feel free to comment after any of the questions if the available answer options do not make sense or do not cover your view in its entirety.

Case load size

1. Psychiatric case load can be measured by breaking down direct patient care activities into new assessments, follow up of stable patients; follow up of unstable patients and emergencies. Strongly Agree ☐	Agree ☐ Neither Agree nor Disagree ☐ Disagree ☐ Strongly disagree ☐
2. Teams value and prefer CCP to be more available rather than see lots of cases. Strongly Agree ☐	Agree ☐ Neither Agree nor Disagree ☐ Disagree ☐ Strongly disagree ☐
3. It is not viable for CCPs to have consultative and strategic roles as well as large clinical caseload

Strongly Agree ☐	Agree ☐ Neither Agree nor Disagree ☐ Disagree ☐ Strongly disagree ☐

4. I think my case load is more than my non-medical colleagues in my team.

Yes ☐ No ☐ Not Sure ☐

5. I think CCPs are the backstop for clinical work where others are not available. Yes ☐ No ☐ Not Sure ☐
6. I think a CCP holds the ultimate responsibility for both individual clients and the availability of resources Yes ☐ No ☐ Not Sure ☐
7. Some CCPs consider a Large case load a “badge of honour”

Yes ☐ No ☐ Not Sure ☐

8. My case load on an average is (If working less than full time please give an approximation of how much the case load would be if you were working full time)

<50 ☐, 50-100 ☐ 100-150 ☐ 150-200 ☐ >200 ☐

9. Complex cases form ________ percentage of my work load

Challenges/effects of large case loads

1. Bigger case load does not leave enough time to provide consultation and respond to urgent requests for assessment	Strongly Agree ☐	Agree ☐ Neither Agree nor Disagree ☐ Disagree ☐ Strongly disagree ☐
2. Large caseloads eat into my Continued Professional Development time	Strongly Agree ☐	Agree ☐ Neither Agree nor Disagree ☐ Disagree ☐ Strongly disagree ☐

3. Bigger case loads may lead to burnout (a combination of emotional exhaustion, depersonalisation and low personal accomplishment)	Strongly Agree ☐	Agree ☐ Neither Agree nor Disagree ☐ Disagree ☐ Strongly disagree ☐
4. Not having special interest cases may lead to burn out. Strongly Agree ☐	Agree ☐ Neither Agree nor Disagree ☐ Disagree ☐ Strongly disagree ☐
5. The high workload and work stress can affect the health of a CCP and thus lead to a negative impact on the provision of patient care. Strongly Agree ☐	Agree ☐ Neither Agree nor Disagree ☐ Disagree ☐ Strongly disagree ☐

Strategies used by CCPs

1. Case Load management CLM (review of case-loads to allow CCP to focus on a smaller number of complex cases while having flexible time available to respond to crises and support other team members) is an important part of a CCPs clinical practice. Strongly Agree ☐	Agree ☐ Neither Agree nor Disagree ☐ Disagree ☐ Strongly disagree ☐

2. CLM was part of my training curriculum as a trainee	Yes ☐ No ☐ Not Sure ☐
3. CLM is part of the training that I provide to my Trainees Yes ☐ No ☐ Not Applicable ☐
4. I feel there is a need to proactively manage my Case Load Yes ☐ No ☐ Not Sure ☐
5. I had decided the size of my case load before I started my CCP post Yes ☐ No ☐ Do not want to comment ☐
6. My case load size is not under my control but is determined by service need	Strongly Agree ☐	Agree ☐ Neither Agree nor Disagree ☐ Disagree ☐ Strongly disagree ☐
7. My case load size and complexity is dependent on the skill mix of other clinicians in my team. Strongly Agree ☐	Agree ☐ Neither Agree nor Disagree ☐ Disagree ☐ Strongly disagree ☐
8. The CCPs case load size and complexity is dependent on the ratio of CCP to non-medical clinicians in the team	Strongly Agree ☐	Agree ☐ Neither Agree nor Disagree ☐ Disagree ☐ Strongly disagree ☐
9. I think each clinician should audit his/her time in order to review their way of working and increase their clinical efficiency	Strongly Agree ☐	Agree ☐ Neither Agree nor Disagree ☐ Disagree ☐ Strongly disagree ☐

10. I think Psychiatrists should have specific training in CLM. Strongly Agree ☐	Agree ☐ Neither Agree nor Disagree ☐ Disagree ☐ Strongly disagree ☐
11. I know when my case load is too Large	Yes ☐ No ☐ Not Sure ☐
12. I know my Case Load is too Large when ….
13. I decide on the number of cases and complexity of the cases to take on my case load by doing the following-
14. I manage my case load by doing the following :
15. The CLM guidelines/tools which help me decide my case load number and complexity and/or help guide my CLM are:

New Ways Of Working (NWW)

1. Do you use NWW in your clinical practice (In clinical practice it will look like - smaller caseloads with additional responsibility of supervising and supporting staff, learning and development, research, service improvement, etc. And /or See patients when needed rather than routinely and working directly with a smaller number of the most complex cases while providing advice and consultancy support to team members, primary care and other partners on a larger scale)

Yes ☐ No ☐ Not Sure ☐

*If you answered No to Q1 then Please* *Click here* *and complete the Consent at the end of the form*

2. Has NWW helped you manage your case load more effectively compared to traditional ways of working?

Yes ☐ No ☐ Not sure ☐

3. If you answered yes to the above please comment on how NWW helped?

*Consent: I would like to take part in a 20 minute one to one interview with the Investigator to discuss the above responses in detail?* **YES ☐ NO ☐ ____________________________________________________________________________________**

*End of the Questionnaire –Thank you very much for taking out time and sharing your experience with me.*
